# Rapid Utilization of Telehealth in a Comprehensive Cancer Center as a Response to COVID-19

**DOI:** 10.1101/2020.04.10.20061259

**Authors:** Peter E. Lonergan, Samuel L. Washington, Linda Branagan, Nathaniel Gleason, Raj S. Pruthi, Peter R. Carroll, Anobel Y. Odisho

**Affiliations:** Department of Urology, Helen Diller Family Comprehensive Cancer Center, University of California, San Francisco, California, USA; Telehealth Resource Center, University of California, San Francisco, California, USA; Division of General Internal Medicine, University of California, San Francisco, California, USA; Center for Digital Health Innovation, University of California, San Francisco, California, USA

**Keywords:** telehealth, COVID-19, video visit, oncology

## Abstract

**Background:** The emergence of the coronavirus disease 2019 (COVID-19) pandemic in March 2020 created unprecedented challenges in the provision of scheduled ambulatory cancer care. As a result, there has been a renewed focus on video consultations as a means to continue ambulatory care.

**Objective:** To analyze the change in video visit volume at the University of California, San Francisco (UCSF) Comprehensive Cancer Center in response to COVID-19 and compare demographics/appointment data from January 1, 2020 and in the 11 weeks after transition to video visits.

**Methods:** Patient demographics and appointment data (dates, visit types, and departments) were abstracted from the Electronic Health Record reporting database. Video visits were performed using a HIPAA-compliant video conferencing platform with a pre-existing workflow.

**Results:** In 17 departments and divisions at the UCSF Cancer Center, 2,284 video visits were performed in the 11 weeks before COVID-19 changes with an average (SD) of 208 (75) per week and 12,946 video visits were performed in the 11 week post-COVID-19 period with an average (SD) of 1,177 (120) per week. The proportion of video visits increased from 7-18% to 54-72%, between the pre- and post-COVID-19 periods without any disparity based on race/ethnicity, primary language, or payor.

**Conclusions:** In a remarkably brief period of time, we rapidly scaled the utilization of telehealth in response to COVID-19 and maintained access to complex oncologic care at a time of social distancing.

## Introduction

The emergence of the coronavirus disease 2019 (COVID-19) pandemic in the United States in March 2020 created unprecedented challenges in the provision of scheduled healthcare and, in particular, ambulatory cancer care. The rapid spread of COVID-19 has renewed focus on telehealth [1], including video consultations [2], as a means of continuing ambulatory care without increasing the risk of potential exposure for patients, clinicians, and staff.

Telehealth is the provision of healthcare remotely by means of a variety of telecommunication platforms such as messaging, audio, and video [3]. While the use of telehealth to deliver cancer care is not new [4] and has already been well described [5], delivering it at the current scale as a result of the COVID-19 pandemic is unprecedented.

The University of California, San Francisco (UCSF) Health established a Telehealth Program in 2015 and offers video visits in all practices. In response to the evolving pandemic, leadership challenged the organization to transition all in-person clinic visits, beginning March 15, 2020, to video visits with exceptions only for specific, urgent cases.

In this study, we analyze the change in video visit volume at the UCSF Comprehensive Cancer Center in response to COVID-19 and compare demographics/appointment data from January 1, 2020 and in the 11 weeks after transition to video visits.

## Methods

Demographics and appointment data (dates, visit types, and departments) were abstracted from the Electronic Health Record reporting database. The pre-COVID-19 period was defined as the 11 weeks from January 1 to March 13, 2020, prior to the transition, and the post-COVID-19 period as the 11 weeks following the transition to video visits on March 16, 2020 up to May 31, 2020. All video visits were performed using a HIPAA-compliant video conferencing platform (Zoom Video Communications Inc, San Jose, CA) with a pre-existing workflow (Figure 1). R version 3.5.3, was used for analysis [6] and a P value less than 0.05 was considered significant.

**Figure 1.**
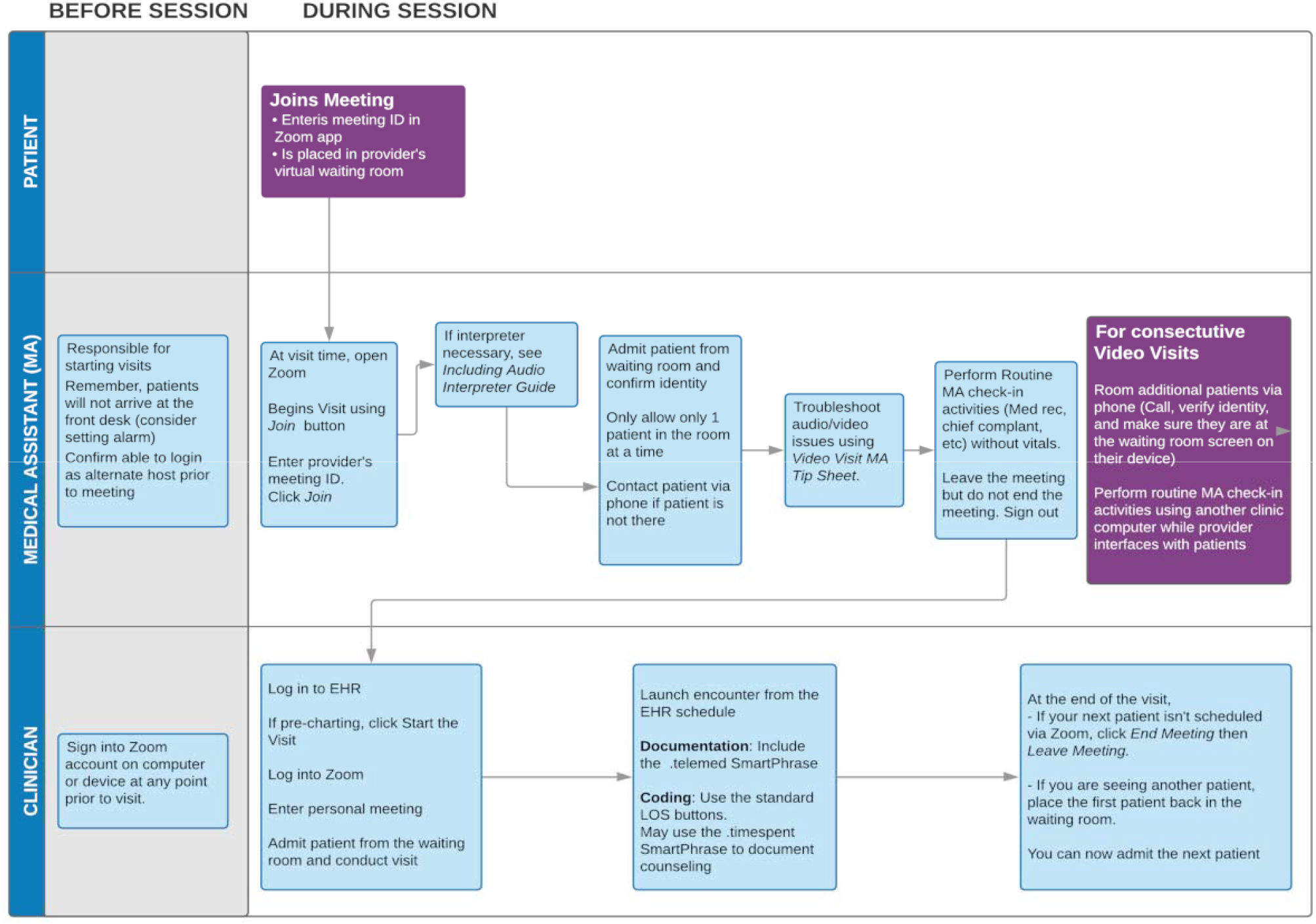
Video visit workflow. Workflow used at the University of California, San Francisco for video visits.

## Results

In the pre-COVID-19 period from January 1 to March 13, 2020, there were a total of 23,988 ambulatory care episodes, with an average (SD) of 2,181 (522) per week across 17 departments and divisions at the UCSF Cancer Center (Figure 2). During this period, a total 2,284 video visits were performed with an average (SD) of 208 (75) video visits being performed per week. The proportion of video visits ranged from 7-18% whereas the proportion of in-person visits ranged from 76-86% (Figure 3). In the post-COVID-19 period from March 16 to May 31, 2020, there was a total of 20,567 ambulatory care episodes, with an average (SD) of 1,870 (200) per week. A total of 12,946 video visits were performed in the post-COVID-19 period, with an average (SD) of 1,177 (120) per week. The proportion of video visits increased to 54-72%. The proportion of episodes during which a procedure was performed ranged from 4-7% in the pre-COVID-19 period and 1-5% in the post-COVID-19 period.

**Figure 2.**
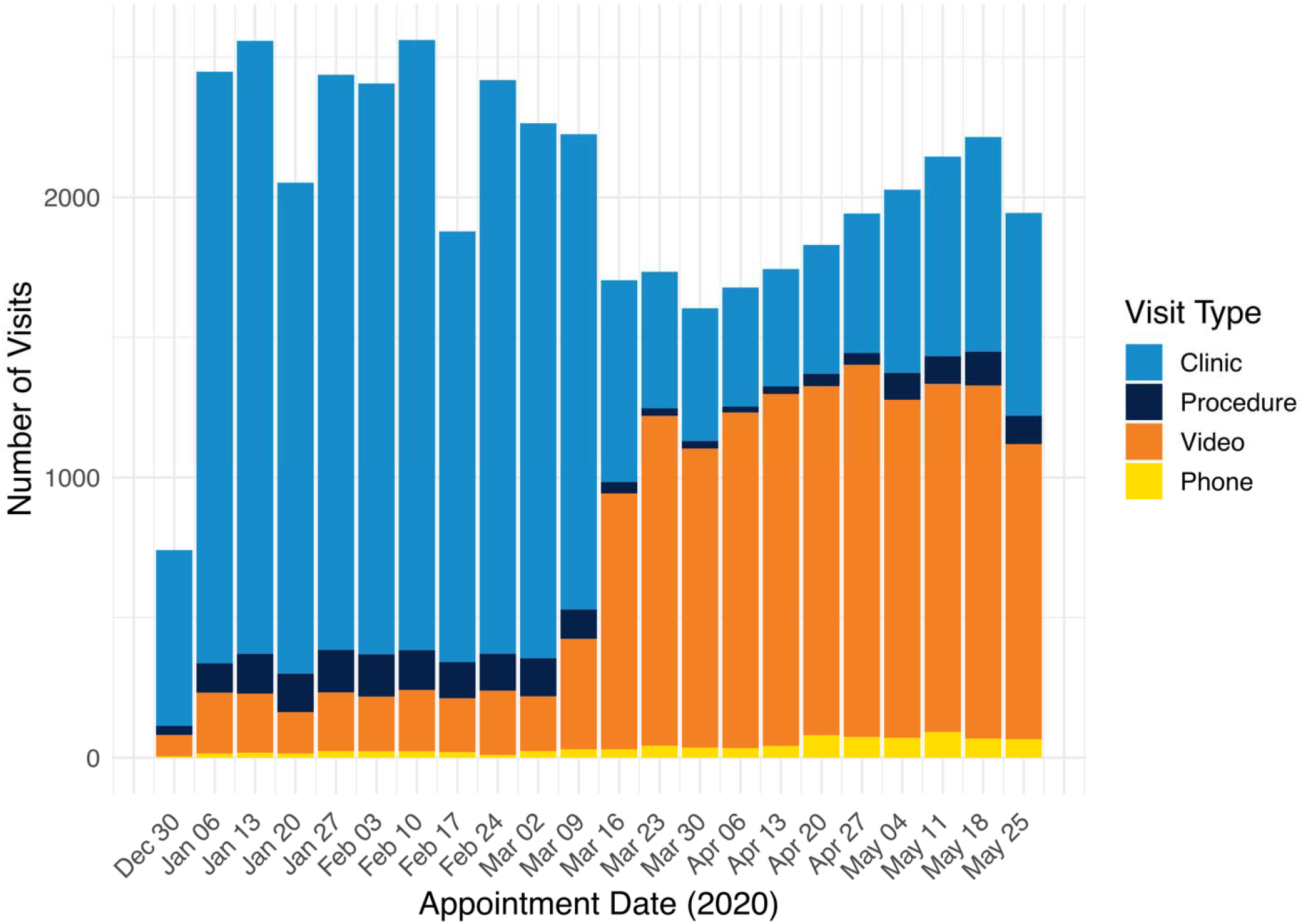
Number of clinic encounters stratified by visit type. The number of in-person visits, procedural visits, video visits and phone visits from January 1 to May 31, 2020 with March 16, 2020 denoting the institution-wide transition to video visits in response to COVID-19.

**Figure 3.**
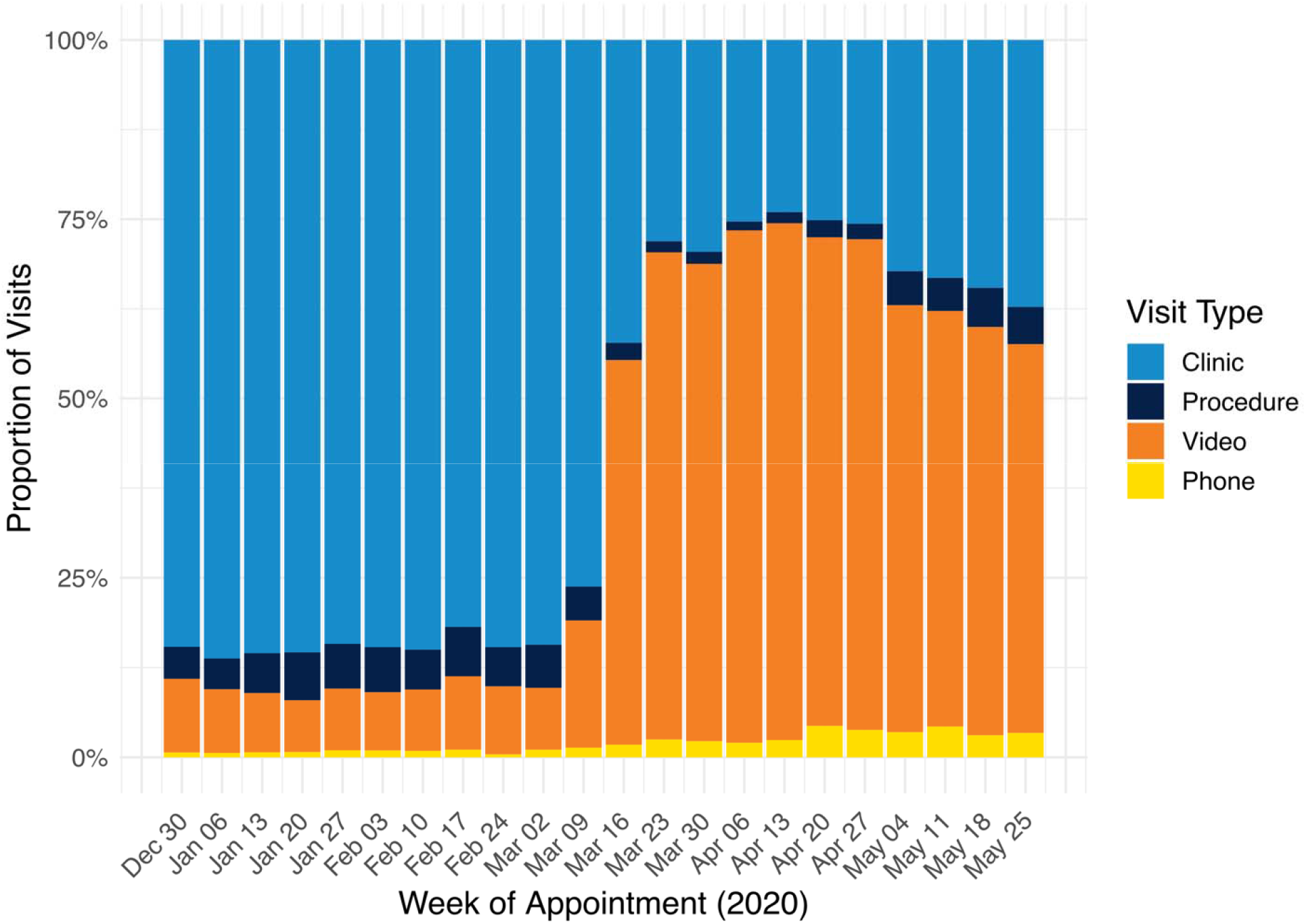
Relative weekly trends in clinic encounters, stratified by visit type. The proportion of in-person visits, procedural visits, video visits and phone visits from January 1 to May 31, 2020 with March 16, 2020 denoting the institution-wide transition to video visits in response to COVID-19.

Table 1 shoes the demographic data of the 2,284 patients who had a video visit in the pre-COVID-19 period and the 12,946 patients who had a video visit in the post-COVID-19 period. In the post COVID-19 period, more Black/African Americans (4.1% vs 3.4%; p<0.001), Hispanic/Latino (11.2% vs 9.4%; p<0.001) and Asians (14.7% vs 8.6%; p<0.001) received care via video visits compared to the pre-COVID-19 period. There was increased post-COVID-19 utilization of video visits for patients in urban areas (92.8% vs 88.7%; p<0.001). We did not find any difference in the insurance status of patients using video visits during either period. In the post COVID-19 period, first clinic encounter (24.9% vs 18.3%; p <0.001) and MD-provided visits (81.8% vs 67.3%; p<0.001) increased.

**Table 1.**
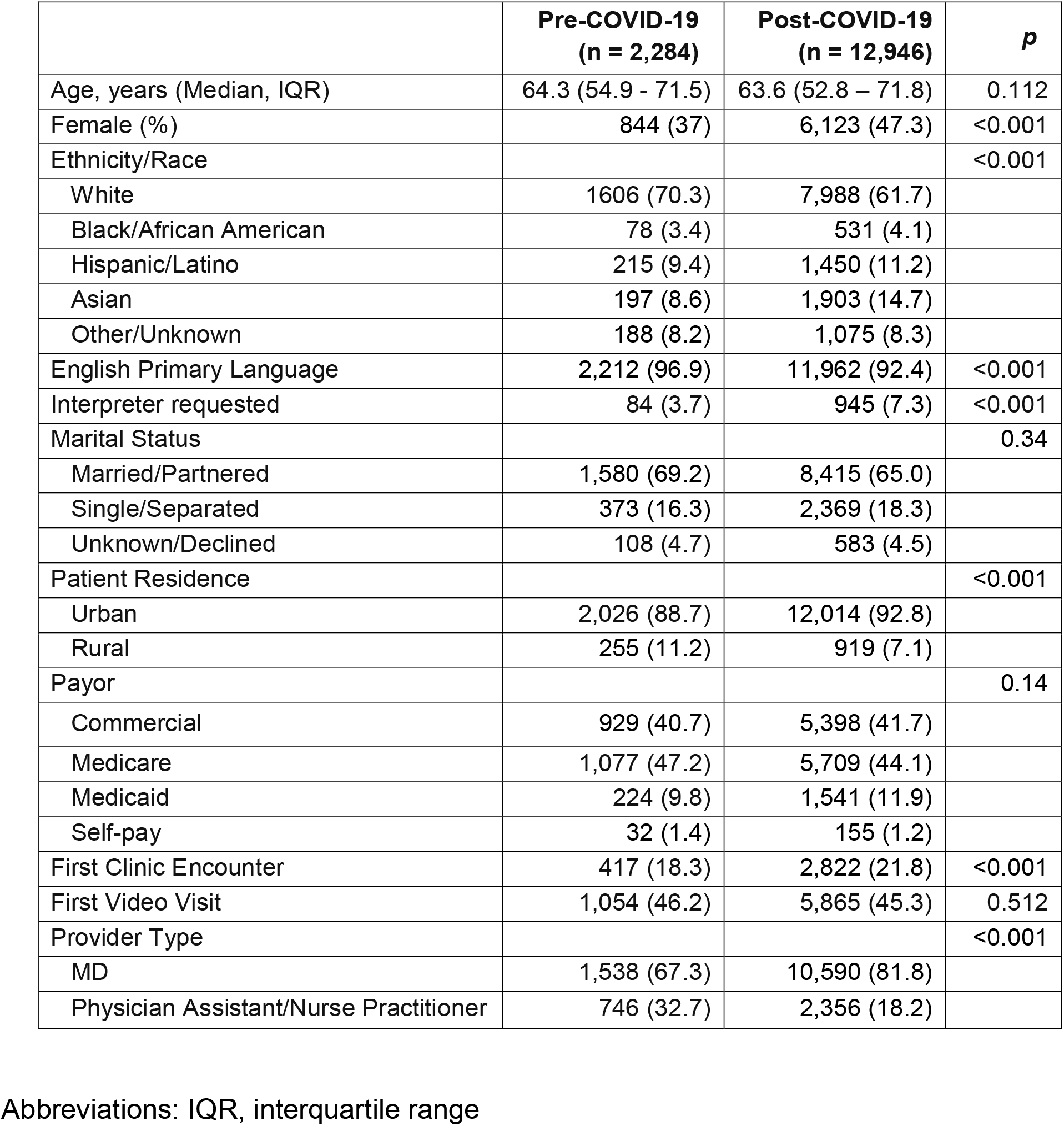
Demographics of patients who had video visits pre- and post-COVID-19

## Discussion

We demonstrate a rapid expansion (from <20% to 72%) in telehealth use in a Comprehensive Cancer Center over a remarkably brief time period in response to COVID-19 without differences by race or insurance. Medicare telehealth visits have increased by more than 25% annually for the past decade [7], yet absolute adoption numbers remain low and fragmented with concerns about potentiating disparities in healthcare access [8]. The vast majority of cancer care cannot be delayed and the COVID-19 pandemic has presented new challenges that telehealth is uniquely situated to solve. Changes that would typically encompass months of planning, pilot testing, and education have been compressed into days. The use of telehealth has grown exponentially with some practices transitioning to near-complete virtual care in as little as a few days [9-11].

Several factors have likely enabled the rapid expansion of video visits at our institution. First, we had an established telehealth structure and workflow familiar to providers and practice staff. Second, UCSF made a strategic decision to provide Relative Value Unit (wRVU) credit to providers for telehealth visits since 2015, irrespective of payer reimbursement. Third, new regulatory changes at federal and state level as a response to COVID-19 have reduced barriers, including the ability to see new patients (including Medicare beneficiaries) without a prior in-person visit to establish care and reimbursement for telehealth encounters by the Centers for Medicare & Medicaid Services (CMS) at parity with in-person visits beginning March 17, 2020 [12]. Finally, CMS now permits providers licensed in any state to provide telehealth services across the country [13].

There are a number of limitations of this study which need to be acknowledged. We did not evaluate the outcome of the video visits including patient satisfaction, qualitative or clinical outcomes. The study is from a single, large urban academic cancer center in the US and our findings may not be generalizable to other specialties, practices or locations. We believe this is the first report of the utilization of video visits in a Comprehensive Cancer Center in response to COVID-19 with a detailed description of the changing demographic of patients utilizing video visits before and after the COVID-19 pandemic.

## Conclusions

Overall, the proportion of video visits increased from 7-18% to 54-72%, between the pre- and post-COVID-19 periods whilst maintaining access to complex oncologic care at a time of social distancing. The COVID-19 pandemic has forced us to radically rethink and change our cancer care delivery models. In many health systems, there will undoubtedly be many lessons learned from this “natural experiment”, which has the potential to permanently change care delivery patterns.

## Data Availability

On request

## Author Contributions

Dr. Odisho had full access to all of the data in the study and takes responsibility for the integrity of the data and the accuracy of the data analysis.

Concept and design: Odisho, Lonergan, Washington.

Acquisition, analysis, or interpretation of data: Odisho, Lonergan, Washington. Drafting of the manuscript: Lonergan, Washington.

Critical revision of the manuscript for important intellectual content: All authors.

Statistical analysis: Odisho, Washington, Lonergan

Administrative, technical, or material support: Branagan. Supervision: Carroll, Pruthi, Odisho

## Additional Contributions

None

